# Epidemiology of malaria and district-level factors associated with malaria elimination in Sumatra region, Indonesia: a retrospective analysis of surveillance data

**DOI:** 10.1101/2025.08.28.25333185

**Authors:** Karina Dian Lestari, Henry Surendra, Bimandra Adiputra Djaafara, Ihsan Fadilah, Lenny L. Ekawati, Hermawan Susanto, Hellen Dewi Prameswari, Sri Budi Fajariyani, Dedy Supriyanto, Herdiana Herdiana, Eamon Conway, Pimphen Charoen, Saranath Lawpoolsri, Wirichada Pan-ngum, Iqbal Elyazar

## Abstract

**Background:** As transmission declines, malaria is likely to become more spatially and demographically concentrated. This study investigated individual-level malaria characteristics and district-level factors associated with elimination in Sumatra, Indonesia.

**Methods:** We analysed individual-level data of 13,386 reported malaria cases from 2019 to 2021. Logistic regression was used to examine individual-level factors associated with imported cases, and district-level socioeconomic and environmental factors associated with elimination. Spatial clustering of indigenous cases was assessed using Global Moran’s *I* and SaTScan.

**Findings:** Among reported cases, 71.8% (9,617) were *P. vivax*, 18.4% (2,467) *P. falciparum,* and 2.8% (369) probable *P. knowlesi*. Of 11,390 investigated cases, 8,748 (76.8%) were indigenous, 1,741 (15.3%) imported, and 901 (7.9%) unclassified. Imported cases were associated with working age, males (aOR 1.73; 95% CI 1.47-2.04), working in mobile and migrant population-related occupations (aOR 1.21; 95% CI 1.01-1.43), diagnosed at hospitals or clinics (aOR 1.31; 95% CI 1.06-1.61), infected with *P. falciparum* (aOR 1.39; 95% CI 1.16-1.67), and hospitalised (aOR 19.77; 95% CI 16.48-23.77). At district level, higher human development index (aOR 1.15; 95% CI 1.01-1.34), lower historical annual parasite incidence (aOR 0.46; 95% CI 0.29-0.69), and fewer endemic neighbours (aOR 0.78; 95% CI 0.67-0.89) were associated with a higher likelihood of elimination. Spatial analysis revealed clustering tendencies, with localised clusters for *P. falciparum*, *P. vivax*, and probable *P. knowlesi*.

**Interpretation:** Imported malaria, local clusters, and socioeconomic and environmental factors affecting the elimination progress. This highlights the need for strengthened surveillance and targeted interventions.

**Funding:** Australian DFAT under SPARK project and WHO Indonesia under TSA of MOTION project.

## Background

Malaria is a vector-borne disease that remains one of the main global health issues, particularly in low- and middle-income countries. WHO reported that the global number of estimated malaria cases has increased from 236 million in 2019 to 263 million in 2023. Despite substantial efforts and control strategies, progress towards elimination has stalled and is currently off track. Key challenges include funding shortfalls, the spread of drug and insecticide resistance, and the impact of climate change on vector distribution and transmission dynamics.^1^ Furthermore, the emergence of zoonotic *P. knowlesi* infection^2^ and inequalities in healthcare access^3^ further complicate malaria control and elimination efforts.

Indonesia has been one of the major malaria hotspots in Southeast Asia region, with an estimated 1.1 million cases and 1.9 thousand deaths according to WHO.^1^ The National Malaria Program (NMP) reported that 401 of 514 districts had been declared malaria-free, leaving about 7% of Indonesia’s population (21 million people) in low to moderate risk of malaria infections.^4^ However, despite a considerable reduction in malaria incidence over the past decade, Indonesia’s goal for achieving malaria elimination in 2030 still encounters challenges. The decreasing trend has slowed since 2014, and resurgences of cases were also observed.^5^ Eliminating malaria in Indonesia is hindered by challenges of limited health systems capacity, uneven distribution of healthcare facility, the complexity of detecting and controlling multi-species infections, and the difficulty of preventing resurgence driven by imported malaria cases.^6^

Sumatra is the largest island of Indonesia that is facing complex challenges of eliminating malaria. For examples, the emergence of *P. knowlesi* infections in humans was recently confirmed by studies conducted in Aceh and North Sumatra provinces.^7,8^ NMP has also identified difficulties in reducing malaria among Suku Anak Dalam, an indigenous communities in Jambi province.^6^ Moreover, malaria-free districts in Sumatra are at risk of malaria importation and reintroduction due to the high movement of the mobile and migrant population (MMP), including forest workers, miners, and agricultural workers, which often engage in outdoor and overnight activities carried out in malaria-endemic areas.

While some district in Sumatra have successfully interrupted transmission, others continue to sustain malaria, reflecting substantial heterogeneity. It also has been known that as malaria transmission rates decrease, the distribution of cases becomes more concentrated, clustering within populations that share similar social, behavioural, and geographical risk factors.^9^ Therefore, identifying and targeting these clusters of malaria cases becomes imperative for a more effective and efficient malaria control.^10^ This study aimed to describe epidemiological characteristics of reported malaria cases, investigate individual-level factors associated with malaria imported cases, identify indigenous malaria clusters, and explore district-level socioeconomic and environmental factors associated with higher likelihood of malaria elimination across 154 districts in Sumatra.

## Methods

### Study setting

Sumatra is the largest island in Indonesia and is home to 21.72% of the national population, totalling to 59 million people according to the 2020 census. The island spans an area of 480,793.28 km^2^ and has a population density of 123 people per km^2^. Administratively, Sumatra is divided into 10 provinces, 154 districts, 1,961 sub-districts, and 25,511 villages. The region is served by 2,683 primary health centres (PHCs) and 752 hospitals, with at least one PHC available in every sub-district (administrative level 3).^11^

### Study design and data sources

This study was a longitudinal retrospective study utilising individual-level national malaria surveillance data recorded in the SISMAL v2 (Malaria Information System) from 2019 to 2021. The positive cases were confirmed by microscopy or rapid diagnostic test (RDT), which can come from passive case detection (PCD) or active case detection (ACD). All *P. knowlesi* cases were reported as probable *P. knowlesi* since polymerase chain reaction (PCR) confirmation is rarely performed, and diagnosis were based on microscopy. Cases were classified as indigenous if they had no travel history to a malaria-endemic region, and as imported if they had a travel history to a malaria-endemic region within the last 2-4 weeks, based on the epidemiological investigation. If a malaria-free district had a single case of indigenous *P. falciparum* or *P. vivax*, the district was classified as having an outbreak. Occupation categories were simplified into two main groups: 1) MMP-related occupations (fisherman, farmer, miner, police/military, plantation worker, forest worker, fish farmer, and blue-collar labourers), and 2) non-MMP-related occupations (housewife, employee, merchant, student, entrepreneur, unemployed, and others). Cases below 15 years old could only be categories as student (non-MMP) in the occupation. List of variables available from SISMAL v2 and used in the analysis was summarised in Table 1.

**Table 1.** Summary of variables available in SISMAL v2 and used in analysis.

|  | Variables |
| --- | --- |
| Demographic | Age group |
|  | Gender |
|  | Pregnancy status |
|  | Occupation |
| Diagnostic and treatment | Health facility type |
|  | Case finding method |
|  | Diagnostic method |
|  | Parasite species |
|  | Treatment |
| Severity | Severity degree |
|  | Hospitalisation status |
|  | Outcome |
| Epidemiological investigation | Case classification |
|  | Source of infection |
|  | Malaria recurrent status |
|  | Primaquine 14 days completion |
|  | Suspected location of infection |

Population data at the health facility level were collected from SISMAL v2. If the PHC did not have population data, we used the subdistrict population data from the Districts in Figures reports by the Indonesian Central Bureau of Statistics. District-level risk factors data in 2021, such as the human development index (HDI), percentage of agricultural workers, and percentage of rural village data, were collected from the Indonesia Central Bureau of Statistics. Annual Parasite Incidence (API) data at the district level from 2010 to 2018 were obtained from the SISMAL v1, and the numbers of neighbouring endemic districts were obtained by calculating the district adjacency matrix using the queen contiguity, which is that the districts would be considered as neighbours if they share a common edge and vertex. Land use data, including forest cover, cropland and flooded vegetation, and settlement areas, were obtained from the ESA Sentinel-2 Satellite^12^ at 10-meters resolution and aggregated to the district level.

### Statistical analysis

The descriptive characteristics of malaria cases were analysed by summarising the incidence and calculating the proportion across the three-year study period. Bivariable and multivariable logistic regression analyses were conducted to explore the individual-level characteristics associated with imported cases. The multivariable analysis included only cases that undergone epidemiological investigation, which enable the classification of imported and indigenous cases. A complete case analysis was employed for these analyses.

At the district level, logistic regression was used to investigate the association between socioeconomic and environmental factors with malaria elimination status. This analysis looked at malaria elimination status in Sumatra, comprising 36 endemic districts (codes as 0) and 106 malaria-free districts (coded as 1), at the end of the study period in 2021. Explanatory factors included land usage, HDI, percentage of agricultural workers, percentage of urban villages, API in 2010, and the number of neighbouring endemic districts.

Before running the multivariable regression, bivariable regression was employed first to identify potential variables, with those showing a p-value <0.10 included in the multivariable model. Multicollinearity was assessed using Variance Inflation Factor (VIF), and variable with a VIF values greater than five was removed. This process was repeated until all remaining variables had VIF values close to one, indicating no collinearity. Backwards stepwise was then performed to select the most important variables. Finally, the model with the lowest AIC was selected. The odds ratio (OR) and 95% confidence interval (CI) were summarised in a table, with p-values less than 0.05 considered statistically significant. The analysis was done using R 4.4.1.

### Spatial heterogeneity of indigenous malaria

The spatial relationship across the study area was first assessed using Global Moran’s *I*. Kulldorff’s spatial scan statistics were then performed using SaTScan™ v10.1.2 to identify clusters of indigenous malaria at the PHC level. Cases recorded at hospitals were reassigned to the nearest PHCs, as hospitals do not have a definitive population. The analysis tested the null hypothesis that the risk of malaria incidence within the population inside a cluster was the same as outside. The high-risk malaria clusters were detected in purely spatial settings using circular window, with a maximum cluster size set at 50% population to avoid pre-selection bias and to allow detection of small and large clusters. Under the Poisson assumption, the likelihood ratio for each circle is calculated by:

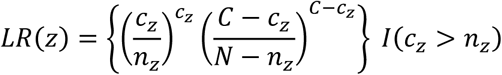

Where 𝑍 is the collection of health facilities in the study region and zone 𝑧 consists of neighbouring geographic units with varying sizes. Then 𝑐_𝑧_ is the observed number of cases and 𝑛_𝑧_is the expected number of cases. Subsequently, 𝐶 = ∑_𝑧_ 𝑐_𝑧_ is the total number of cases and 𝑁 = ∑_𝑧_ 𝑛_𝑧_ is the total number of expected cases. The indicator function of 𝐼 ( ) will be equal to 1 when the cluster has more cases than expected cases and 0 otherwise.

The most likely cluster, referred to as the primary cluster in this paper, was defined as the one with the maximum log-likelihood ratio (LLR), assessed at a statistical significance level of 𝛼 = 0.05 using 999 Monte Carlo replications. Once the primary cluster was identified, SaTScan removed it and re-ran the analysis to detect additional, non-overlapping clusters, referred to as secondary clusters. The Gini coefficient was used as a more systematic method than using an arbitrary number of maximum spatial window sizes to report the significant clusters.^13^ Cluster map was edited using QGIS version 3.34.5.

### Ethics

Permission to access and use the malaria surveillance data was granted by the Malaria Working Group, Ministry of Health, Jakarta, Indonesia. Ethical approval for this study was obtained from the Ethics Committee of the Faculty of Tropical Medicine, Mahidol University (Submission No. TMEC 24-001).

## Results

### Epidemiological characteristics

Between 2019 and 2021, a total of 13,386 malaria cases were reported across 127 districts in Sumatra, while 27 districts reported zero cases. The highest number of cases (4,722) occurred in 2021. Most cases were detected in PHCs (88.4%) through passive case detection (83.8%) and diagnosed using microscopy (57.9%). *P. vivax* accounted for 71.8% (9,617) of the total cases, while *P. falciparum* and probable *P. knowlesi* comprised 18.4% (2,467) and 2.8% (369), respectively. Female cases accounted for 36.3%, with only 3.1% of these cases being pregnant. Nearly half of the cases (48.2%) occurred in adults aged 25-64, with a considerable portion seen in young adults aged 15-24 (24.2%) and children aged under 15 years old (25.3%). Approximately 20% of patients required hospitalisation. Severe malaria cases and malaria-related deaths were rare, with 1% severe cases and a total of fourteen deaths. The completeness of data varied across variables, with missing values ranging from 0.3% for age group to 20.9% for completion of primaquine (PQ) medication (Table 2).

**Table 2.** Summary of malaria incidence in Sumatra region.

|  | 2019 | 2020 | 2021 | Total |
| --- | --- | --- | --- | --- |
| Number of districts reporting malaria cases | 104 (67.5) | 88 (57.1) | 77 (50.0) | 127 (82.5) |
| Total case | 4,467 | 4,197 | 4,722 | 13,386 |
| **Demography** |  |  |  |  |
| Age group |  |  |  |  |
| < 15 | 1,214 (27.2) | 1,078 (25.7) | 1,095 (23.2) | 3,387 (25.3) |
| 15-24 | 991 (22.2) | 1,051 (25.0) | 1,194 (25.3) | 3,236 (24.2) |
| 25-64 | 2,129 (47.7) | 1,979 (47.2) | 2,346 (49.7) | 6,454 (48.2) |
| 65+ | 95 (2.1) | 89 (2.1) | 87 (1.8) | 271 (2.0) |
| Unknown | 38 (0.9) | 0 (0) | 0 (0) | 38 (0.3) |
| Gender |  |  |  |  |
| Female | 1,623 (36.3) | 1,569 (37.4) | 1,668 (35.3) | 4,860 (36.3) |
| Male | 2,842 (63.6) | 2,628 (62.6) | 3,054 (64.7) | 8,524 (63.7) |
| Unknown | 2 (0) | 0 (0) | 0 (0) | 2 (0) |
| Pregnancy status |  |  |  |  |
| Pregnant | 61 (3.8) | 49 (3.1) | 41 (2.5) | 151 (3.1) |
| Not pregnant | 1,488 (91.7) | 1,461 (93.1) | 1,541 (92.4) | 4,490 (92.4) |
| Unknown | 74 (4.6) | 59 (3.8) | 86 (5.2) | 219 (4.5) |
| Occupation |  |  |  |  |
| Non-MMP | 2,953 (66.1) | 2,777 (66.2) | 3,297 (69.8) | 9,027 (67.4) |
| MMP | 1,385 (31.0) | 1,346 (32.1) | 1,334 (28.3) | 4,065 (30.4) |
| Unknown | 129 (2.9) | 74 (1.8) | 91 (1.9) | 294 (2.2) |
| **Diagnostic and treatment** |  |  |  |  |
| Health facility type |  |  |  |  |
| PHC | 3,730 (83.5) | 3,895 (92.8) | 4,209 (89.1) | 11,834 (88.4) |
| Hospital/Clinic | 737 (16.5) | 302 (7.2) | 513 (10.9) | 1,552 (11.6) |
| Case detection method |  |  |  |  |
| ACD | 554 (12.4) | 689 (16.4) | 823 (17.4) | 2,066 (15.4) |
| PCD | 3,859 (86.4) | 3,482 (83.0) | 3,878 (82.1) | 11,219 (83.8) |
| Unknown | 54 (1.2) | 26 (0.6) | 21 (0.4) | 101 (0.8) |
| Diagnostic method |  |  |  |  |
| Microscopy | 2,952 (66.1) | 2,536 (60.4) | 2,265 (48.0) | 7,753 (57.9) |
| RDT | 1,492 (33.4) | 1,636 (39.0) | 2,447 (51.8) | 5,575 (41.6) |
| PCR | 9 (0.2) | 9 (0.2) | 1 (0) | 19 (0.1) |
| Unknown | 14 (0.3) | 16 (0.4) | 9 (0.2) | 39 (0.3) |
| Parasite species |  |  |  |  |
| *P. vivax* | 2,480 (55.5) | 3,190 (76.0) | 3,947 (83.6) | 9,617 (71.8) |
| *P. falciparum* | 1,496 (33.5) | 564 (13.4) | 407 (8.6) | 2,467 (18.4) |
| Mix | 346 (7.7) | 246 (5.9) | 257 (5.4) | 849 (6.3) |
| Probable *P. knowlesi* | 101 (2.3) | 174 (4.1) | 94 (2.0) | 369 (2.8) |
| Other | 4 (0.1) | 3 (0.1) | 8 (0.2) | 15 (0.1) |
| Unknown | 40 (0.9) | 20 (0.5) | 9 (0.2) | 69 (0.5) |
| Treatment |  |  |  |  |
| Standard treatment | 4,240 (94.9) | 4,024 (95.9) | 4,519 (95.7) | 12,783 (95.5) |
| Non-standard treatment | 7 (0.2) | 4 (0.1) | 2 (0) | 13 (0.1) |
| Unknown | 220 (4.9) | 169 (4.0) | 201 (4.3) | 590 (4.4) |
| **Severity** |  |  |  |  |
| Hospitalisation |  |  |  |  |
| Inpatient | 1,315 (29.4) | 671 (16.0) | 744 (15.8) | 2,730 (20.4) |
| Outpatient | 2,975 (66.6) | 3,484 (83.0) | 3,910 (82.8) | 10,369 (77.5) |
| Unknown | 177 (4.0) | 42 (1.0) | 68 (1.4) | 287 (2.1) |
| Severity degree |  |  |  |  |
| Severe malaria | 45 (1.0) | 21 (0.5) | 64 (1.4) | 130 (1.0) |
| Uncomplicated malaria | 4,306 (96.4) | 4,156 (99.0) | 4,644 (98.3) | 13,106 (97.9) |
| Unknown | 116 (2.6) | 20 (0.5) | 14 (0.3) | 150 (1.1) |
| Outcome |  |  |  |  |
| Death | 10 (0.2) | 2 (0) | 2 (0) | 14 (0.1) |
| Recovered | 4,055 (90.8) | 3,925 (93.5) | 4,574 (96.9) | 12,554 (93.8) |
| Unknown | 402 (9.0) | 270 (6.4) | 146 (3.1) | 818 (6.1) |
| **Epidemiological investigation** |  |  |  |  |
| Case classification |  |  |  |  |
| Investigated | 3,012 (67.4) | 3,710 (88.4) | 4,668 (98.9) | 11,390 (85.1) |
| Not investigated | 1,455 (32.6) | 487 (11.6) | 54 (1.1) | 1,996 (14.9) |
| Source of infection |  |  |  |  |
| Indigenous | 1,967 (65.3) | 2,872 (77.4) | 3,909 (83.7) | 8,748 (65.4) |
| Imported | 694 (23.0) | 408 (11.0) | 639 (13.7) | 1,741 (13.0) |
| Unknown | 351 (11.7) | 430 (11.6) | 120 (2.6) | 901 (6.7) |
| Malaria recurrent status * |  |  |  |  |
| Relapse case | 154 (9.0) | 178 (6.0) | 436 (11.1) | 768 (9.0) |
| New case | 1,286 (75.3) | 1,598 (54.3) | 3,430 (87.3) | 6,314 (73.6) |
| Unknown | 267 (15.6) | 1,168 (39.7) | 62 (1.6) | 1,497 (17.4) |
| PQ completion † |  |  |  |  |
| Complete | 1,146 (55.8) | 2,876 (90.2) | 3,216 (76.8) | 7,238 (76.8) |
| Incomplete | 52 (2.5) | 11 (0.3) | 17 (0.4) | 80 (0.8) |
| Unobserved | 104 (5.1) | 32 (1.0) | 7 (0.2) | 143 (1.5) |
| Unknown | 751 (36.6) | 271 (8.5) | 945 (22.6) | 1,967 (20.9) |
| Suspected origin |  |  |  |  |
| Sumatera | 168 (24.2) | 148 (36.3) | 348 (54.5) | 664 (38.1) |
| Java & Bali | 3 (0.4) | 0 (0) | 0 (0) | 3 (0.2) |
| Papua | 209 (30.1) | 18 (4.4) | 66 (10.3) | 293 (16.8) |
| Unknown | 314 (45.2) | 242 (59.3) | 225 (35.2) | 781 (44.9) |

Data in n (%). MMP = Mobile and migrant population. PHC = Primary health centre. ACD = Active case detection. PCD = Passive case detection. RDT = Rapid diagnostic test. PCR = Polymerase chain reaction. PQ = Primaquine.
\* Malaria recurrent status was defined from investigated *P. vivax* cases.
† PQ medication was considered complete if the patients take all PQ for 14-days. PQ completions were calculated from investigated *P. vivax* and Mix cases.

A total of 11,390 cases (85.1%) were investigated, of which 8,748 (76.8%) were classified as indigenous, 1,741 (15.3%) as imported, and 901 (7.9%) remained unclassified. Interestingly, indigenous cases were reported in 27 malaria-free districts, indicating outbreak events. Rokan Hilir district in Riau province recorded the highest number of outbreak cases, accounting for 20% of all cases (2,675/13,386). The outbreaks peaked in June and October 2020.

Indigenous and imported cases exhibited different epidemiological characteristics based on the bivariable and multivariable logistic regression analyses (n = 10,163), except for treatment received and malaria outcomes (Table 3). After adjusting for other variables, imported cases were associated with working age groups, being male (aOR 1.73; 95% CI 1.47-2.04), working in MMP-related occupation (aOR 1.21; 95% CI 1.01-1.43), diagnosed at hospitals or clinics (aOR 1.31; 95% CI 1.06-1.61), infected with *P. falciparum* (aOR 1.39; 95% CI 1.16-1.67), and being hospitalised (aOR 19.77; 95% CI 16.48-23.77).

**Table 3.**
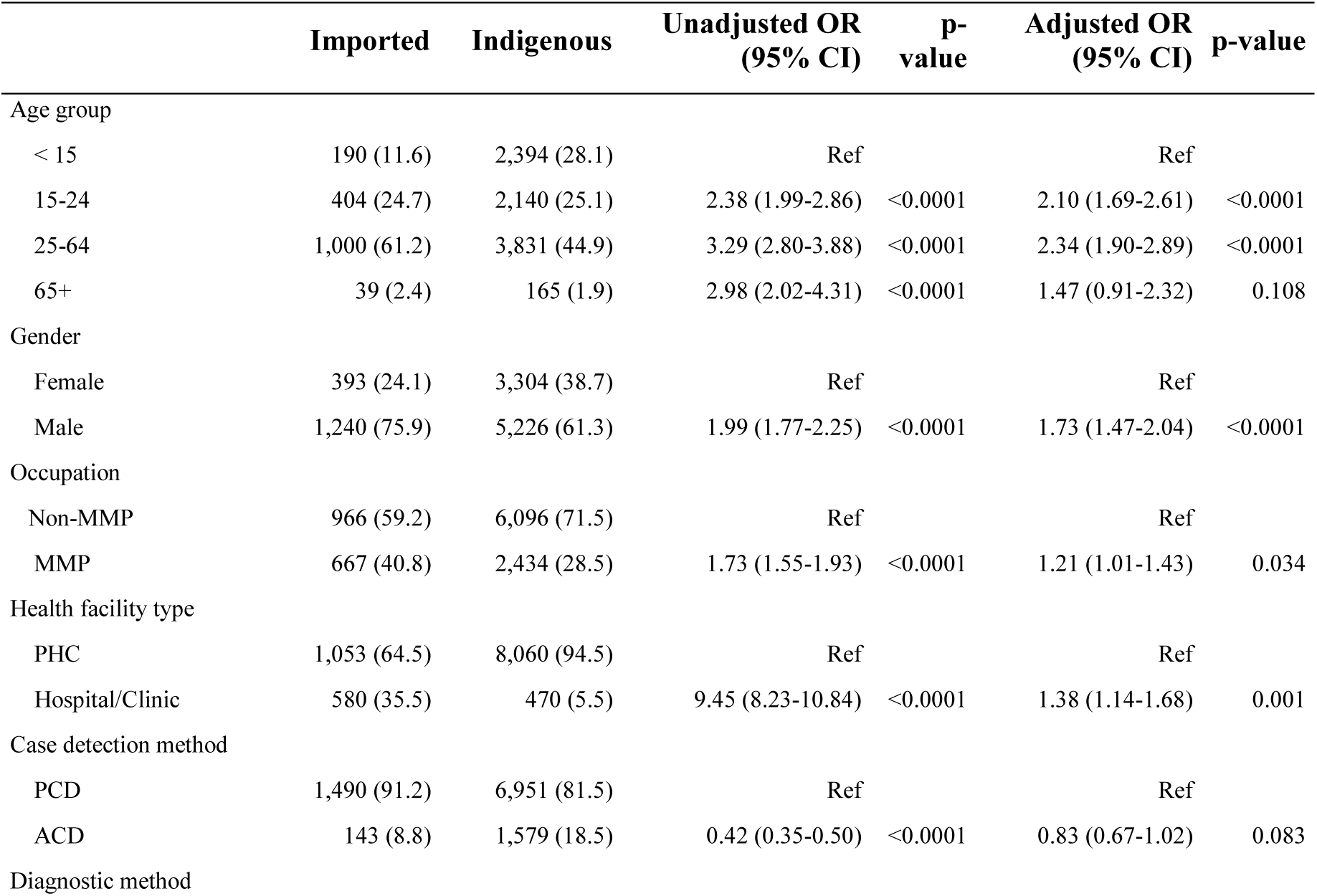
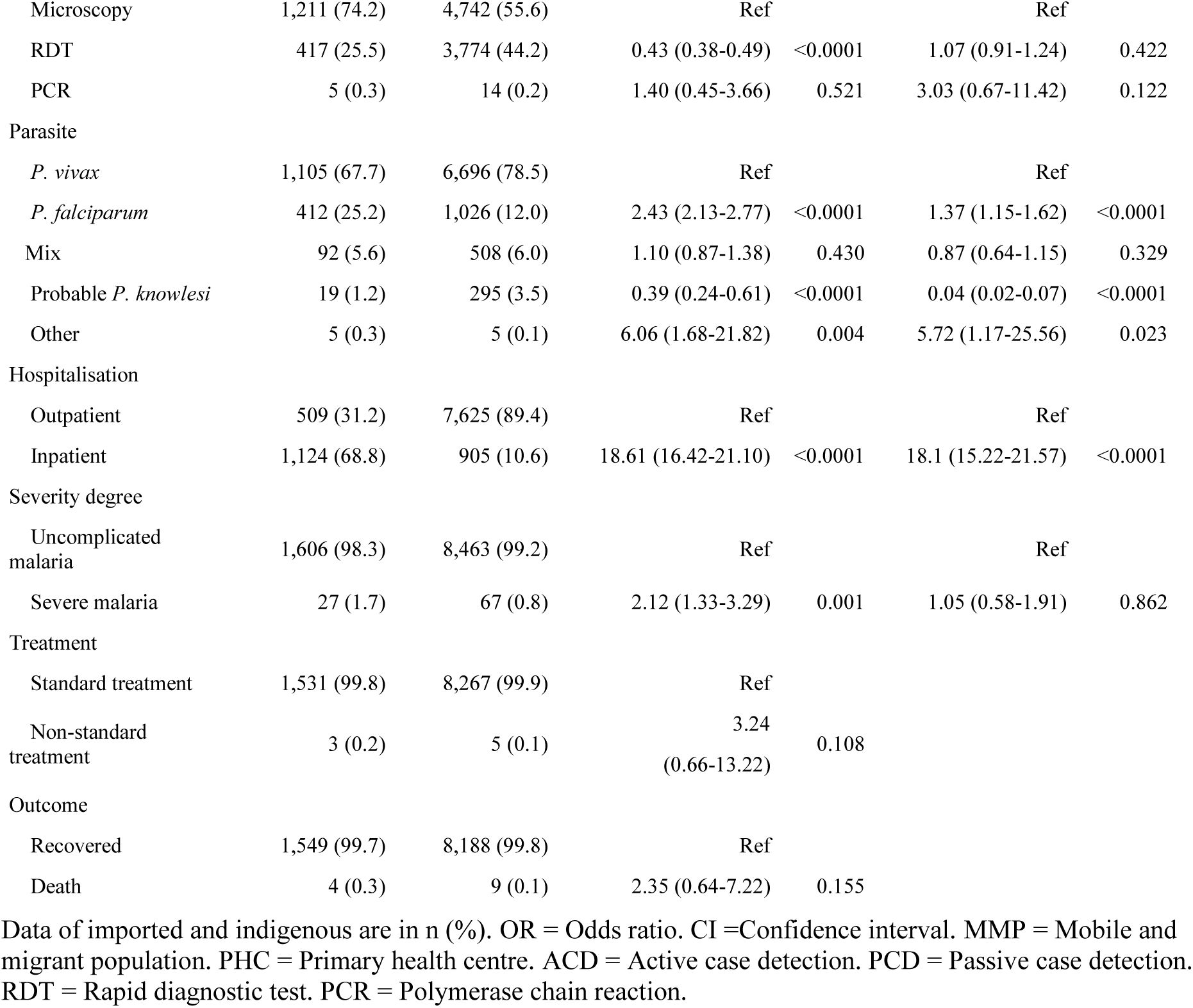
Comparison of characteristic between imported and indigenous cases. Using n = 10,163.

We were able to determine the suspected origin of the importation for around half of the investigated cases (960; 55.1%). Most of the importations originated from other districts in Sumatra (38.1%) and Papua (16.8%). Almost half of the imported cases (48.6%) came from low-endemic districts, and around 20% were from very high-endemic districts (API ≥ 100). Notably, 93 imported cases (9.7%) originated from malaria-free districts. Flow maps indicated that the movement of imported cases, both from outside (Figure 1) and within (Figure 2) Sumatra, fluctuated yearly, with 2020 being the lowest due to the COVID-19 mobility restriction.

**Figure 1.**
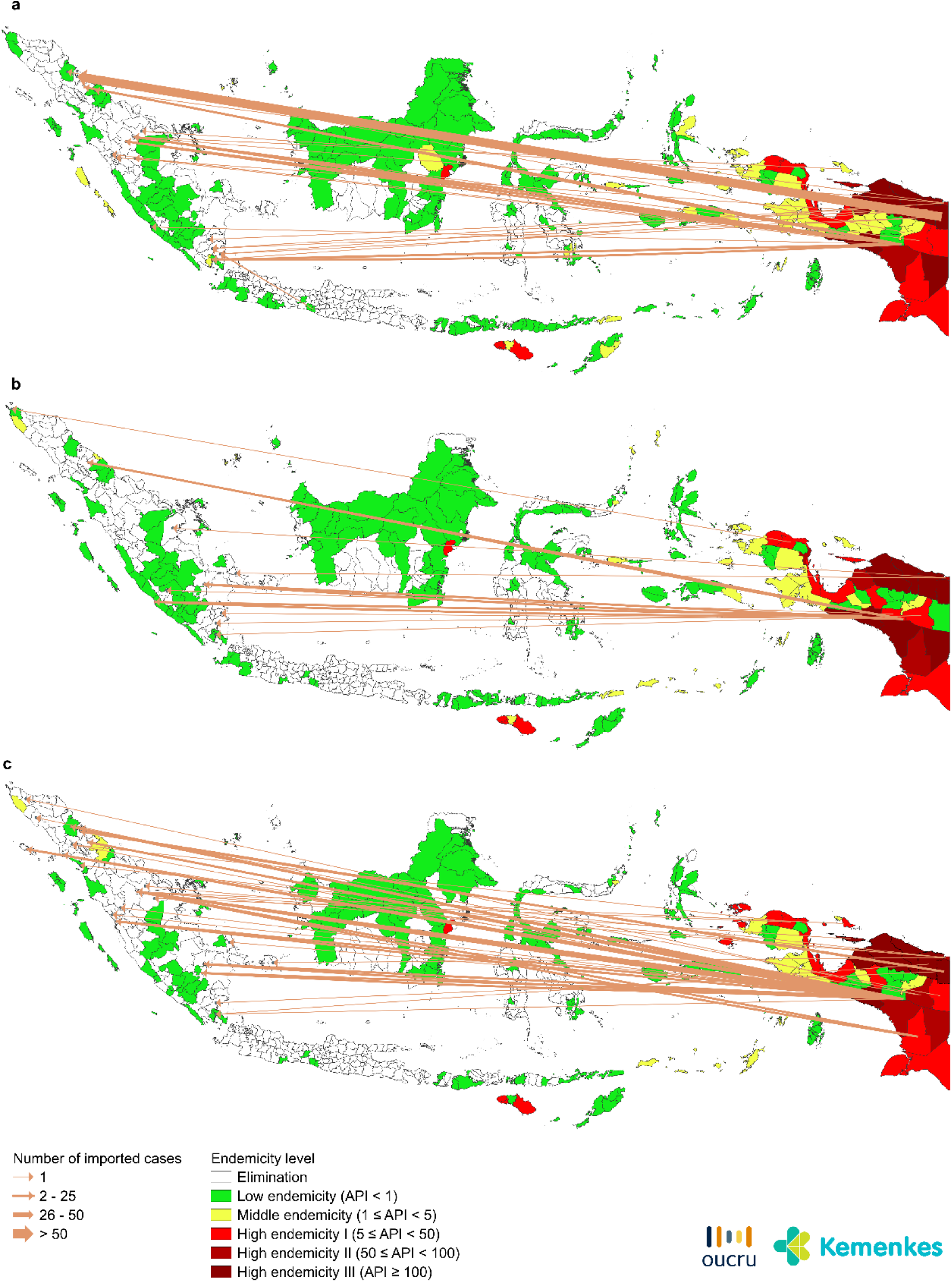
Flow map of imported malaria cases from outside Sumatra region, a) 2019, b) 2020, c) 2021.

**Figure 2.**
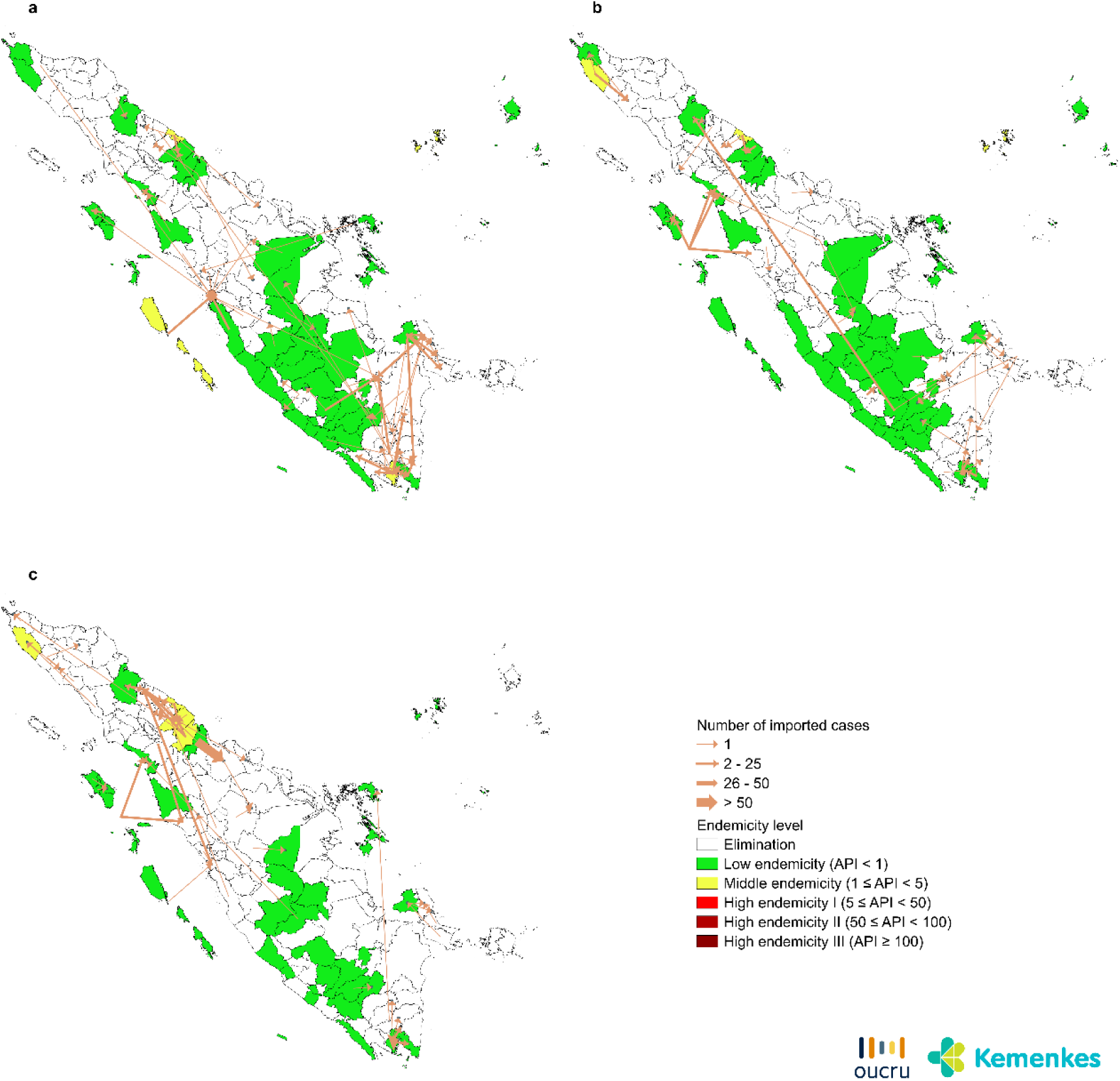
Flow map of imported malaria cases from within Sumatra region, a) 2019, b) 2020, c) 2021.

### District-level socioeconomic and environmental factors associated with malaria elimination

Multivariable logistic regression shows that the human development index, API in 2010, and the number of neighbouring endemic districts were significantly associated with districts achieving malaria elimination (Table 4). The higher the HDI is, the more likely it is for the district to eliminate malaria (aOR 1.15; 95% CI 1.01-1.34). On the contrary, a higher historical API in 2010 made it less likely to eliminate malaria (aOR 0.46; 95% CI 0.29-0.69). Similarly, a higher number of neighbouring endemic districts decreased the likelihood of malaria elimination (aOR 0.78; 95% CI 0.67-0.89).

**Table 4.**
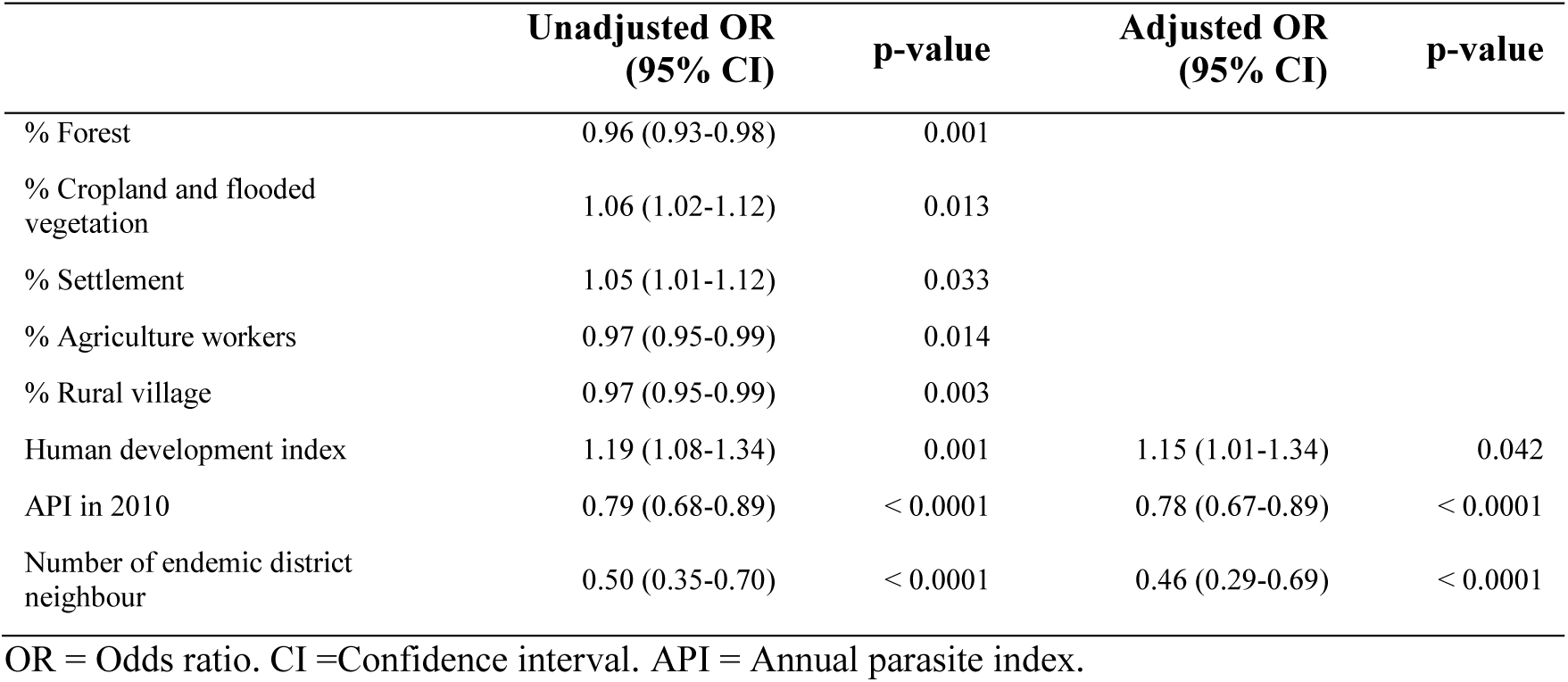
Socioeconomic and environmental factors associated with malaria elimination.

|  | Unadjusted OR<br>(95% CI) | p-value | Adjusted OR<br>(95% CI) | p-value |
| --- | --- | --- | --- | --- |
| % Forest | 0.96 (0.93-0.98) | 0.001 |  |  |
| % Cropland and flooded vegetation | 1.06 (1.02-1.12) | 0.013 |  |  |
| % Settlement | 1.05 (1.01-1.12) | 0.033 |  |  |
| % Agriculture workers | 0.97 (0.95-0.99) | 0.014 |  |  |
| % Rural village | 0.97 (0.95-0.99) | 0.003 |  |  |
| Human development index | 1.19 (1.08-1.34) | 0.001 | 1.15 (1.01-1.34) | 0.042 |
| API in 2010 | 0.79 (0.68-0.89) | < 0.0001 | 0.78 (0.67-0.89) | < 0.0001 |
| Number of endemic district neighbour | 0.50 (0.35-0.70) | < 0.0001 | 0.46 (0.29-0.69) | < 0.0001 |

OR = Odds ratio. CI =Confidence interval. API = Annual parasite index.

### Spatial heterogeneity of indigenous malaria

The number of PHC used for the indigenous cluster analysis was 2,460, with 9,271 cases, representing 69% of the total cases in Sumatra from 71 districts. Spatial autocorrelation analysis using Global Moran’s *I* showed significant clustering of malaria cases across the region (Moran’s *I* value = 0.285, z score = 4.485, p-value < 0.001). The primary clusters of indigenous malaria cases were detected in different provinces each year for each *P. falciparum* and *P. vivax* (Figure 3 and Figure 4, respectively), except for the probable *P. knowlesi* cluster (Figure 5), which was confined to Aceh province. No overlapping clusters were observed between species. Several secondary clusters were also detected, with *P. vivax* having the largest number of clusters (Supplementary 1).

**Figure 3.**
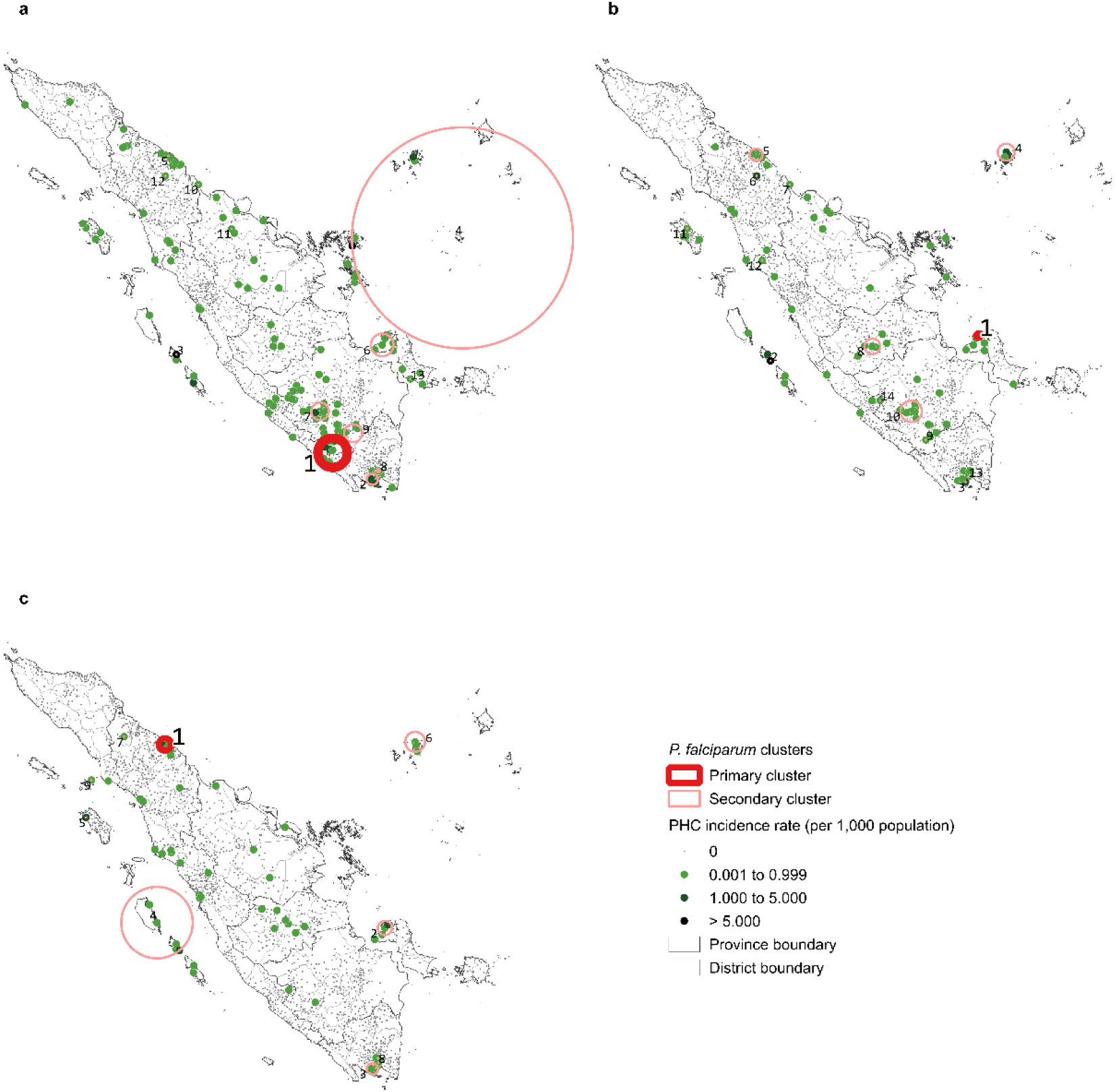
Clusters of *P. falciparum* in a) 2019, b) 2020, c) 2021.

**Figure 4.**
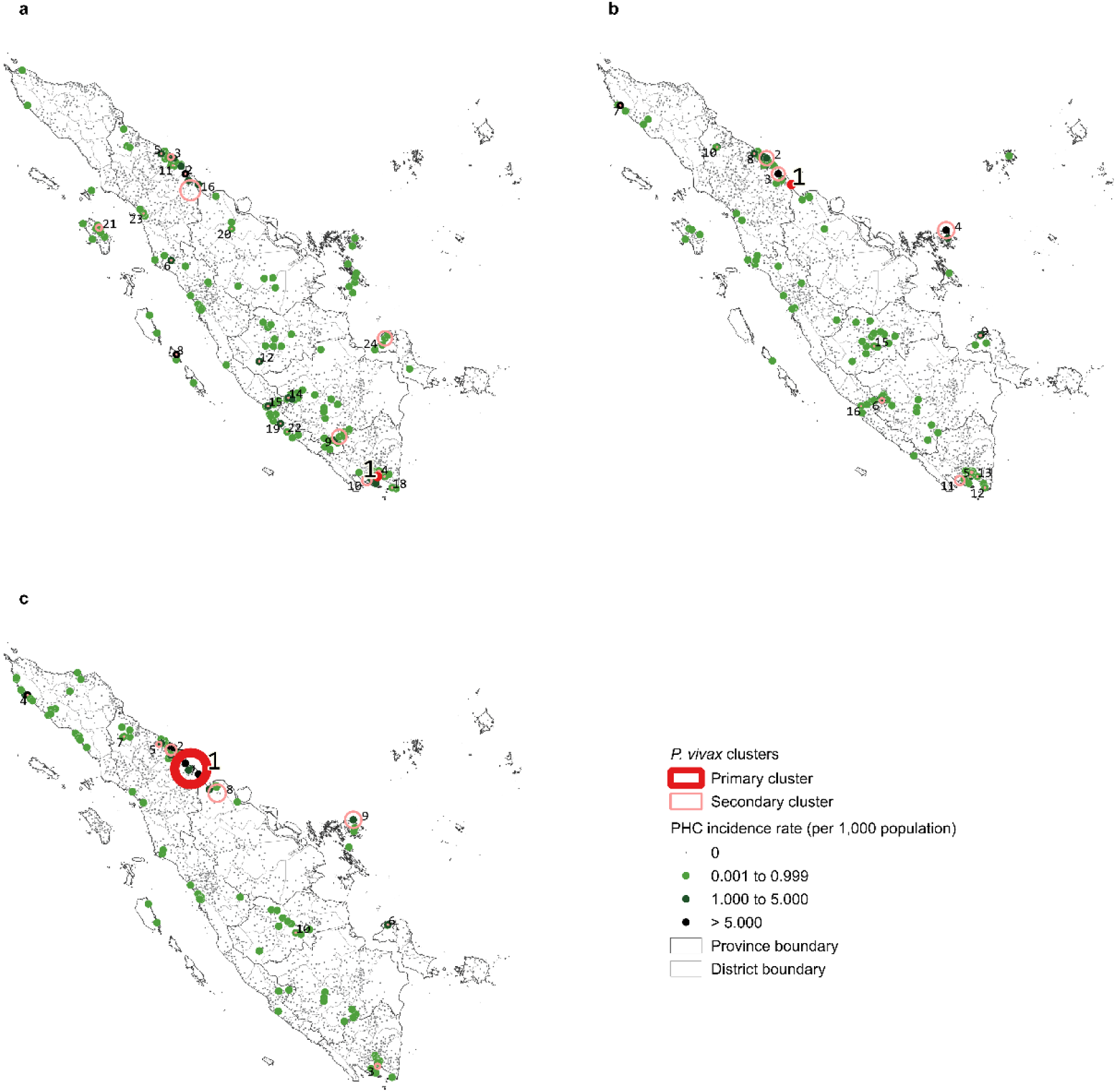
Clusters of *P. vivax* in a) 2019, b) 2020, c) 2021.

**Figure 5.**
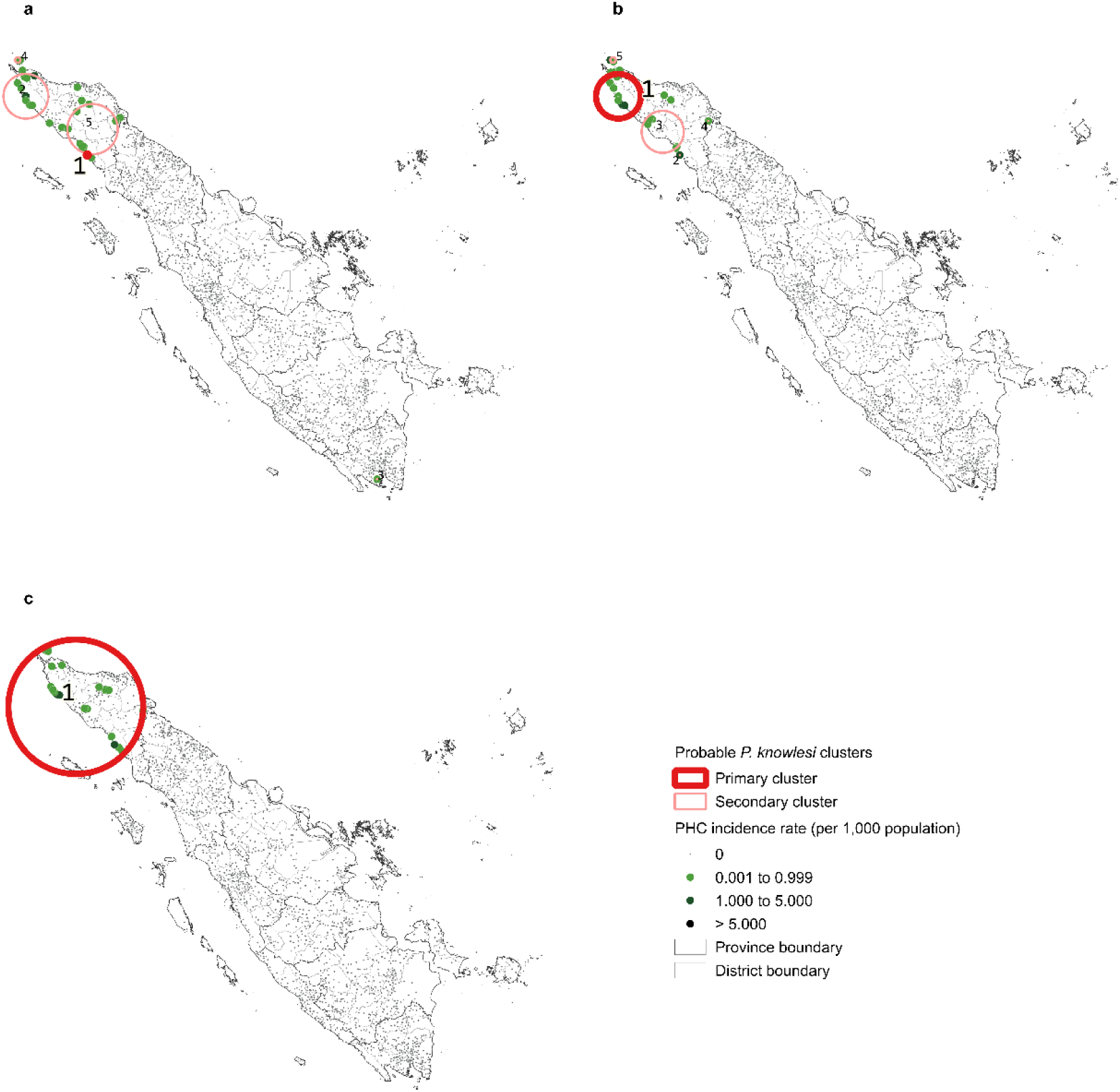
Clusters of probable *P. knowlesi* in a) 2019, b) 2020, c) 2021.

The Hanura PHC in Lampung province was consistently detected as a *P. vivax* cluster centre for three years. In 2019, as a primary cluster, with an estimated relative risk of 333.4 covering a radius of less than 1 km and a population of 37,400. While in following years, it was detected as a secondary cluster. In 2020, the primary cluster for *P. vivax* was centred at Panipahan PHC, Riau province, which had a radius of less than 1 km and a population of 38,402. The estimated risk of malaria within this cluster was 1,563.7 times higher compared to the surrounding areas, indicating the outbreak in the Rokan Hilir district concentrated in this PHC catchment area. In 2021, the cluster centre shifted to the neighbouring Sei Berombang PHC in North Sumatra province, which had a radius of 48.9 km and a population of 418,029. The estimated relative risk of this cluster was 111.1.

## Discussion

This study is among the first to investigate the epidemiology of malaria and district-level socioeconomic and environmental factors associated with the likelihood of malaria elimination across 154 districts in Sumatra, Indonesia. Our analysis revealed that while *P. vivax* infections dominated malaria cases, *P. falciparum* remained prevalent, and a substantial proportion of probable *P. knowlesi* infections was also reported. Around 15% of the total reported cases were imported, contributing to the reintroduction of malaria transmission in areas already achieved or nearing elimination. Compared to indigenous cases, imported cases were more likely to involve males of working age, working in MMP-related occupations, diagnosed at hospitals or clinics, infected with *P. falciparum*, and being hospitalised. At district-level, higher HDI, lower historical API, and fewer endemic district neighbours were associated with higher likelihood to eliminate malaria. Furthermore, our spatial analysis indicated a clustering tendency, with various localised malaria clusters identified for *P. falciparum*, *P. vivax*, and probable *P. knowlesi*.

The demographic characteristics of malaria cases in Sumatra were predominantly males aged 15-64 years. Several studies have observed this epidemiological shift of increasing cases in adult and males as the region progresses towards the pre-elimination phase.^9^ Our individual-level analysis further revealed that imported malaria cases were strongly associated with MMP-related occupations, consistent with findings from other Southeast Asia settings, including Cambodia^14^ and Malaysia.^15^ Imported cases were also reported originating from malaria-free areas, which may be explained by high population mobility between endemic and malaria-free districts, complicating the identification of transmission origin by the PHC staff. It may also due to relapse of *P. vivax* parasites reintroducing infection where suitable vector habitat exist.^16^ Mandatory screening for MMPs before re-entering malaria-free areas needs to be reinforced to improve early detection and treatment to prevent reintroduction and resurgence of local transmission.

Unfortunately, malaria-free districts in Sumatra still experienced outbreaks during 2019-2021, further highlighting the essential needs of maintaining control and elimination efforts. Investigations by the NMP and independent experts have identified the potential source of transmission that caused the outbreak in Rokan Hilir in 2020, which was an imported case involving fishermen who had stayed overnight in an endemic area. There were also several factors contributing to the outbreaks, including delays in response and reporting, logistical challenges in case detection, and issues in program management.^17^ Additionally, the Indonesian government imposed mobility restrictions and shifted healthcare priorities during COVID-19,^18^ which reduced the ability of healthcare workers to conduct outbreak control measures and routine case detection. Consequently, there was a decline in malaria testing and reporting, as well as delay in LLINs distribution and IRS activity.^6^ A mathematical modelling study investigated that the greatest COVID-19 impact on malaria in low- and middle-income countries was the interruption of LLINs distribution, which can lead to a 36% increase in deaths due to malaria.^19^

Spatial analysis results showed that indigenous malaria incidence in the Sumatra region tended to be clustered in close neighbourhoods. SaTScan detected several localised clusters, with different cluster centres of *P. falciparum* and *P. vivax* each year, which indicates a temporal instability of cluster locations. This instability could be due to the human movement^20^ and ecological factors, such as proximity to breeding sites.^21^ For example, in 2021, the *P. vivax* cluster centre was shifted from the outbreak location in Riau province to adjacent endemic area of North Sumatra province. This spatial pattern suggests that the outbreak may have been driven by imported cases originating from the neighbouring area. By contrast, a cluster in Lampung province persisted throughout the study period, indicating that local sociodemographic and environmental factors may sustain the ongoing transmission.^22^ The stability of this cluster also suggests it as a prime location for targeted control interventions. These dynamics highlight the importance of strong surveillance to detect, report, and respond to importation cases early, while also maintaining targeted interventions in endemic areas to reduce the source of importation and prevent reintroduction in malaria-free areas.

Our district-level analysis suggests that malaria elimination was more likely to be with higher HDI, a lower history of API over the last decade, and a lower number of neighbouring endemic districts. According to the WHO definition, elimination requires three consecutive years without indigenous cases, meaning good health system capacity is essential to sustain malaria control and elimination efforts. As suggested by Cohen et al., the median year from effective control measures to reach malaria elimination was 12 years.^23^ This finding further indicates that malaria-free status is vulnerable to its neighbouring epidemiological situation, as human mobility crosses administrative border. This suggests the need of robust cross-border surveillance, which has been shown to be one of the key interventions to reduce malaria incidence.^24^

Parasite-specific challenges further complicates the elimination efforts in Sumatra. *P. vivax* remains the most difficult species to eliminate due to its ability to relapse and poor adherence to PQ treatment. The completion was as low as 33% in malaria-free districts. This situation highlights the need to improve adherence through supervised treatment,^25,26^ or using text message reminders may be a feasible alternative among the MMP where direct supervision is difficult. Where glucose-6-phosphate-dehydrogenase (G6PD) testing is available, changing the medication to tafenoquine (TQ) should also be considered, as it offers a shorter treatment duration.^27^ Meanwhile, *P. knowlesi* is difficult to diagnose using microscopy because of its morphological similarity to other species, and confirmation requires PCR. However, PCR capacity is limited to a few centres in large cities.^28^ The ability of Aceh province to diagnose probable *P. knowlesi* cases is largely due to the presence of expert-level (L1) microscopists who have been specifically trained to recognise *P. knowlesi* morphology in blood smears. Investing in PCR testing is therefore recommended to accurately assess the burden of *P. knowlesi* in areas identified as high-risk for *P. knowlesi* transmission, such as Aceh and North Sumatra province.^29^

There are several limitations that should be considered when interpreting this study’s findings. First, malaria cases reported by SISMAL may suffer from underreporting and cannot be guaranteed to be free from human error. Secondly, the analysis of district-level association was limited to a few socioeconomic and environmental factors, which may not fully account for potential confounders. Finally, the results from SaTScan should be interpreted with caution, as sparse data distributed across large areas may produce inflated relative risk estimates.^30^ While this does not invalidate the location of the clusters, it reduces the reliability of relative risk values. Aggregating data to a coarser level, such as by district, could mitigate this issue, although it risks losing the finer details needed to identify localised clusters within districts.

## Conclusion

This study demonstrated that malaria cases in Sumatra share similar characteristics with those observed in low-to moderate-elimination settings. Imported cases were particularly prevalent among males of working age and engaged in MMP-related occupations. Although imported cases accounted for only a small proportion, they still pose a threat to elimination efforts if not promptly addressed. We also applied spatial analysis to identify clusters of indigenous malaria cases and highlight priority areas for malaria control. The association between socioeconomic and environmental factors and the likelihood of malaria elimination further emphasise the importance of robust cross-border surveillance and targeted intervention. Future studies incorporating more advanced spatial analysis, with adjustment for relevant explanatory variables including demography, intervention coverage, socioeconomic, and environmental factors, would provide further insight of the malaria clustering and its associated risk. Strengthening the quality of surveillance data will also be essential for formulating evidence to inform effective control measures.

## Authors’ contributions

KDL, WP, and IE conceptualised the study. HS, HDP, SBF, and DS were responsible for data collection and provision. KDL and IE curated and verified the data. KDL was responsible for formal analysis, visualisation, and writing the initial draft. HS, BAD, IF, LLE, and HH provided relevant input and additional context. EC, PC, SL, WP, and IE were responsible for study oversight. All authors had critically reviewed the study for intellectual content and had full responsibility for the decision to submit the final manuscript.

## Data sharing statement

The malaria data that support the findings of this study are from the Malaria Working Group, Ministry of Health of Indonesia, but restrictions apply to the availability of these data, which were used under license for this study, and so are not publicly available. Data are however available upon reasonable request and with written approval from the Malaria Working Group, Ministry of Health of Indonesia. Other data were publicly available at Indonesia Central Bureau of Statistics (https://www.bps.go.id/) and ESRI (https://livingatlas.arcgis.com/landcoverexplorer/). Reproducible code for data analysis and visualisation is available at https://github.com/karinadnlstr/malaria-sumatra.

## Data Availability

The malaria data supporting the findings of this study are subject to access restrictions, as they were used under license for the current study and therefore not publicly available. Data are however available upon reasonable request and with written approval from the Malaria Working Group, Ministry of Health of Indonesia. Other data were publicly available at Indonesia Central Bureau of Statistics (https://www.bps.go.id/) and ESRI (https://livingatlas.arcgis.com/landcoverexplorer/). Reproducible code for data analysis and visualisation is available at https://github.com/karinadnlstr/malaria-sumatra.

https://livingatlas.arcgis.com/landcoverexplorer/

## Acknowledgement

This study was conducted as part of master’s programme in Biomedical and Health Informatics at Mahidol University, which was funded by the Australian Department of Foreign Affairs and Trade (DFAT) under the Strengthening Preparedness in the Asia-Pacific Region through Knowledge (SPARK) project. The data collection was supported by WHO Indonesia under Targeted Scenario Analysis (TSA) of the Mapping of Mobile and Migrant Population at the Outdoor Malaria Transmission Setting in Indonesia (MOTION) project. We are grateful to the healthcare staff for supporting malaria surveillance and data collection activities, and to the Malaria Working Group, Ministry of Health of Indonesia, for granting access to the malaria data.

