## Supplementary 1 for "Epidemiology of malaria and district-level factors associated with malaria elimination in Sumatra region, Indonesia: a retrospective analysis of surveillance data"

**Supplementary material**

Table of content

**Table S1 Summary of *P. falciparum* clusters by year and province**

**Table S2 Summary of *P. vivax* clusters by year and province**

**Table S3 Summary of probable *P. knowlesi* clusters by year and province**

**Table S1 Summary of *P. falciparum* clusters by year and province**

| **Year** | **Cluster** | **Province** | **Radius (km)** | **No of PHC** | **Pop** | **No of cases** | **% case** | **RR** | **p-value** |
| --- | --- | --- | --- | --- | --- | --- | --- | --- | --- |
| **2019** | 1 | South Sumatra | 44.8 | 25 | 461,528 | 309 | 22.6 | 32.1 | < 0.001 |
|  | 2 | Lampung | 17.0 | 7 | 154,647 | 233 | 17.1 | 67.8 | < 0.001 |
|  | 3 | West Sumatra | < 1 | 1 | 7,454 | 117 | 8.6 | 642.7 | < 0.001 |
|  | 4 | Riau Island | 336.2 | 26 | 178,506 | 104 | 7.6 | 23.5 | < 0.001 |
|  | 5 | North Sumatra | < 1 | 1 | 33,025 | 57 | 4.2 | 67.4 | < 0.001 |
|  | 6 | Bangka Belitung Island | 34.3 | 7 | 164,374 | 68 | 5 | 16.2 | < 0.001 |
|  | 7 | South Sumatra | 26.6 | 19 | 421,806 | 91 | 6.7 | 8.6 | < 0.001 |
|  | 8 | Lampung | 5.0 | 15 | 505,526 | 63 | 4.6 | 4.8 | < 0.001 |
|  | 9 | Lampung | 27.4 | 18 | 408,694 | 55 | 4 | 5.2 | < 0.001 |
|  | 10 | Riau | < 1 | 1 | 38,402 | 20 | 1.5 | 19.8 | < 0.001 |
|  | 11 | Riau | < 1 | 1 | 22,550 | 16 | 1.2 | 26.9 | < 0.001 |
|  | 12 | North Sumatra | < 1 | 1 | 21,819 | 14 | 1 | 24.3 | < 0.001 |
|  | 13 | Bangka Belitung Island | < 1 | 1 | 8,336 | 7 | 0.5 | 31.6 | < 0.001 |
| **2020** | 1 | Bangka Belitung Island | 1.8 | 2 | 33,306 | 105 | 19.5 | 375.0 | < 0.001 |
|  | 2 | West Sumatra | < 1 | 1 | 2,423 | 37 | 6.9 | 1570.8 | < 0.001 |
|  | 3 | Lampung | < 1 | 1 | 13,645 | 41 | 7.6 | 311.5 | < 0.001 |
|  | 4 | Riau Island | 25.7 | 4 | 29,857 | 38 | 7.1 | 131.1 | < 0.001 |
|  | 5 | North Sumatra | 19.0 | 14 | 377,376 | 66 | 12.3 | 19.0 | < 0.001 |
|  | 6 | North Sumatra | < 1 | 1 | 21,819 | 27 | 5 | 124.8 | < 0.001 |
|  | 7 | Riau | < 1 | 1 | 38,402 | 29 | 5.4 | 76.4 | < 0.001 |
|  | 8 | Jambi | 24.0 | 4 | 97,686 | 19 | 3.5 | 19.3 | < 0.001 |
|  | 9 | South Sumatra | < 1 | 1 | 42,128 | 10 | 1.9 | 23.2 | < 0.001 |
|  | 10 | South Sumatra | 32.5 | 23 | 478,187 | 23 | 4.3 | 4.8 | < 0.001 |
|  | 11 | North Sumatra | < 1 | 1 | 10,141 | 5 | 0.9 | 47.7 | < 0.001 |
|  | 12 | West Sumatra | < 1 | 1 | 37,586 | 7 | 1.3 | 18.1 | 0.001 |
|  | 13 | Lampung | < 1 | 1 | 37,739 | 7 | 1.3 | 18.0 | 0.001 |
|  | 14 | Bengkulu | < 1 | 1 | 12,405 | 5 | 0.9 | 39.0 | 0.002 |
| **2021** | 1 | North Sumatra | 19.0 | 14 | 378,872 | 116 | 33.5 | 68.1 | < 0.001 |
|  | 2 | Bangka Belitung Island | 21.6 | 4 | 77,138 | 56 | 16.2 | 128.9 | < 0.001 |
|  | 3 | Lampung | 17.0 | 7 | 158,466 | 55 | 15.9 | 61.3 | < 0.001 |
|  | 4 | West Sumatra | 108.7 | 10 | 64,950 | 31 | 9 | 78.0 | < 0.001 |
|  | 5 | North Sumatra | < 1 | 1 | 11,774 | 14 | 4 | 184.6 | < 0.001 |
|  | 6 | Riau Island | 30.3 | 5 | 37,966 | 12 | 3.5 | 48.8 | < 0.001 |
|  | 7 | North Sumatra | < 1 | 1 | 16,768 | 6 | 1.7 | 54.2 | < 0.001 |
|  | 8 | Lampung | < 1 | 1 | 26,377 | 5 | 1.4 | 28.6 | 0.009 |
|  | 9 | Aceh | < 1 | 1 | 4,477 | 3 | 0.9 | 100.7 | 0.022 |

Cluster number 1 indicates the most likely cluster. PHC = Primary health centre. RR = Relative risk.

**Table S2 Summary of *P. vivax* clusters by year and province**

| **Year** | **Cluster** | **Province** | **Radius (km)** | **No of PHC** | **Pop** | **No of cases** | **% case** | **RR** | **p-value** |
| --- | --- | --- | --- | --- | --- | --- | --- | --- | --- |
| **2019** | 1 | Lampung | < 1 | 1 | 37,400 | 454 | 19.6 | 333.4 | < 0.001 |
|  | 2 | North Sumatra | < 1 | 1 | 28,921 | 228 | 9.8 | 193.1 | < 0.001 |
|  | 3 | North Sumatra | 9.1 | 4 | 142,092 | 248 | 10.7 | 43.1 | < 0.001 |
|  | 4 | Lampung | 7.6 | 14 | 443,995 | 224 | 9.7 | 12.2 | < 0.001 |
|  | 5 | North Sumatra | < 1 | 1 | 35,059 | 89 | 3.8 | 58.3 | < 0.001 |
|  | 6 | West Sumatra | < 1 | 1 | 37,586 | 84 | 3.6 | 51.2 | < 0.001 |
|  | 7 | Lampung | < 1 | 1 | 36,717 | 83 | 3.6 | 51.8 | < 0.001 |
|  | 8 | West Sumatra | < 1 | 1 | 7,454 | 47 | 2 | 142.2 | < 0.001 |
|  | 9 | South Sumatra | 21.8 | 8 | 215,843 | 114 | 4.9 | 12.2 | < 0.001 |
|  | 10 | Lampung | 14.2 | 3 | 66,106 | 73 | 3.2 | 25.2 | < 0.001 |
|  | 11 | North Sumatra | < 1 | 1 | 35,194 | 39 | 1.7 | 24.9 | < 0.001 |
|  | 12 | Jambi | < 1 | 1 | 16,845 | 31 | 1.3 | 41.2 | < 0.001 |
|  | 13 | Lampung | < 1 | 1 | 22,413 | 31 | 1.3 | 31.0 | < 0.001 |
|  | 14 | Bengkulu | < 1 | 1 | 16,144 | 28 | 1.2 | 38.8 | < 0.001 |
|  | 15 | Bengkulu | < 1 | 1 | 6,352 | 20 | 0.9 | 70.2 | < 0.001 |
|  | 16 | North Sumatra | 30.5 | 12 | 340,621 | 75 | 3.2 | 5.0 | < 0.001 |
|  | 17 | Lampung | < 1 | 1 | 52,685 | 24 | 1 | 10.2 | < 0.001 |
|  | 18 | Lampung | < 1 | 1 | 22,745 | 17 | 0.7 | 16.6 | < 0.001 |
|  | 19 | Bengkulu | < 1 | 1 | 12,747 | 13 | 0.6 | 22.7 | < 0.001 |
|  | 20 | Riau | < 1 | 1 | 22,550 | 15 | 0.6 | 14.8 | < 0.001 |
|  | 21 | North Sumatra | 9.3 | 6 | 128,065 | 30 | 1.3 | 5.2 | < 0.001 |
|  | 22 | Bengkulu | < 1 | 1 | 14,636 | 12 | 0.5 | 18.2 | < 0.001 |
|  | 23 | North Sumatra | < 1 | 1 | 20,470 | 9 | 0.4 | 9.7 | 0.01 |
|  | 24 | Bangka Belitung Island | 21.6 | 4 | 75,318 | 15 | 0.6 | 4.4 | 0.03 |
| **2020** | 1 | Riau | < 1 | 1 | 38,402 | 1693 | 53.8 | 1563.7 | < 0.001 |
|  | 2 | North Sumatra | 21.7 | 13 | 350,867 | 407 | 12.9 | 21.7 | < 0.001 |
|  | 3 | North Sumatra | 19.9 | 2 | 35,691 | 215 | 6.8 | 105.9 | < 0.001 |
|  | 4 | Riau Island | 24.8 | 3 | 20,547 | 147 | 4.7 | 122.9 | < 0.001 |
|  | 5 | Lampung | < 1 | 1 | 37,739 | 129 | 4.1 | 58.4 | < 0.001 |
|  | 6 | Bengkulu | 7.1 | 2 | 20,695 | 72 | 2.3 | 58.3 | < 0.001 |
|  | 7 | Aceh | < 1 | 1 | 9,324 | 51 | 1.6 | 91.1 | < 0.001 |
|  | 8 | North Sumatra | < 1 | 1 | 33,986 | 50 | 1.6 | 24.5 | < 0.001 |
|  | 9 | Bangka Belitung Island | 1.8 | 2 | 33,306 | 24 | 0.8 | 11.9 | < 0.001 |
|  | 10 | North Sumatra | < 1 | 1 | 16,768 | 17 | 0.5 | 16.7 | < 0.001 |
|  | 11 | Lampung | 14.2 | 3 | 66,367 | 28 | 0.9 | 7.0 | < 0.001 |
|  | 12 | Lampung | < 1 | 1 | 22,937 | 17 | 0.5 | 12.2 | < 0.001 |
|  | 13 | Lampung | < 1 | 1 | 37,180 | 17 | 0.5 | 7.5 | < 0.001 |
|  | 14 | Lampung | 2.1 | 3 | 68,367 | 19 | 0.6 | 4.6 | 0.002 |
|  | 15 | Jambi | < 1 | 1 | 17,420 | 10 | 0.3 | 9.4 | 0.003 |
|  | 16 | Bengkulu | < 1 | 1 | 6,352 | 6 | 0.2 | 15.5 | 0.038 |
| **2021** | 1 | North Sumatra | 48.9 | 18 | 418,029 | 1818 | 47.6 | 111.1 | < 0.001 |
|  | 2 | North Sumatra | 16.5 | 16 | 452,675 | 978 | 25.6 | 38.9 | < 0.001 |
|  | 3 | Lampung | 7.9 | 3 | 92,784 | 364 | 9.5 | 58.4 | < 0.001 |
|  | 4 | Aceh | < 1 | 1 | 9,324 | 170 | 4.5 | 257.5 | < 0.001 |
|  | 5 | North Sumatra | 7.8 | 4 | 73,885 | 90 | 2.4 | 16.8 | < 0.001 |
|  | 6 | Bangka Belitung Island | < 1 | 1 | 12,396 | 20 | 0.5 | 21.9 | < 0.001 |
|  | 7 | North Sumatra | < 1 | 1 | 16,768 | 16 | 0.4 | 12.9 | < 0.001 |
|  | 8 | Riau | 26.4 | 6 | 158,267 | 42 | 1.1 | 3.6 | < 0.001 |
|  | 9 | Riau Island | 24.8 | 3 | 21,728 | 16 | 0.4 | 10.0 | < 0.001 |
|  | 10 | Jambi | < 1 | 1 | 17,947 | 10 | 0.3 | 7.5 | 0.02 |

Cluster number 1 indicates the most likely cluster. PHC = Primary health centre. RR = Relative risk.

**Table 3 Summary of probable *P. knowlesi* clusters by year and province**

| **Year** | **Cluster** | **Province** | **Radius (km)** | **No of PHC** | **Pop** | **No of cases** | **% case** | **RR** | **p-value** |
| --- | --- | --- | --- | --- | --- | --- | --- | --- | --- |
| **2019** | 1 | Aceh | < 1 | 1 | 12,262 | 25 | 25 | 1390.1 | < 0.001 |
|  | 2 | Aceh | 67.9 | 32 | 300,539 | 35 | 35 | 91.1 | < 0.001 |
|  | 3 | Lampung | < 1 | 1 | 36,154 | 9 | 9 | 139.8 | < 0.001 |
|  | 4 | Aceh | 8.3 | 4 | 25,512 | 8 | 8 | 174.3 | < 0.001 |
|  | 5 | Aceh | 76.4 | 44 | 442,728 | 16 | 16 | 21.8 | < 0.001 |
| **2020** | 1 | Aceh | 67.9 | 32 | 298,535 | 129 | 75 | 514.9 | < 0.001 |
|  | 2 | Aceh | < 1 | 1 | 13,391 | 18 | 10.5 | 449.7 | < 0.001 |
|  | 3 | Aceh | 63.9 | 36 | 411,918 | 11 | 6.4 | 8.5 | 0.002 |
|  | 4 | Aceh | < 1 | 1 | 18,514 | 4 | 2.3 | 66.3 | 0.004 |
|  | 5 | Aceh | 8.3 | 5 | 33,638 | 4 | 2.3 | 36.5 | 0.025 |
| **2021** | 1 | Aceh | 204.5 | 286 | 4,243,444 | 90 | 100 |  | < 0.001 |

Cluster number 1 indicates the most likely cluster. PHC = Primary health centre. RR = Relative risk.
